# Uromodulin T62P variant causes kidney tubular stress and injury modulated by age and polygenic risk

**DOI:** 10.64898/2026.07.29.26359269

**Authors:** Atlas Khan, Andrea Gresch, Eric Olinger, Marta Mariniello, Ning Shang, Maria Vanessa Perez Gomez, Ian Dinsmore, Holly Mabillard, Tooraj Mirshahi, Alexander R Chang, Olivier Devuyst, Krzysztof Kiryluk

## Abstract

While ultra-rare missense variants in *UMOD* cause highly penetrant autosomal dominant tubulointerstitial kidney disease, a more frequent *UMOD* T62P variant conveys intermediate risk with variable penetrance. To determine whether age or polygenic risk contributes to the variable penetrance of T62P, we combined genotype and phenotype data from 882,306 individuals across the UK Biobank (discovery cohort) and the All of Us and MyCode biobanks (validation cohorts). We also analyzed the impact of aging on uromodulin processing and cellular stress in stably transfected kidney tubular cells expressing wild-type or mutant *UMOD*. We compared the effects of the GPS on risk of CKD between T62P carriers and non-carriers and tested for the GPS-by-T62P interaction. The *UMOD* T62P variant was reproducibly associated with increased risk of CKD in an age-dependent manner. Compared to wild-type, clones of T62P-expressing cells exhibited a defective uromodulin maturation profile, causing endoplasmic reticulum retention and stress. We also observed significant T62P-by-GPS interaction, with T62P carriers in the top quintile of the GPS having over 5-fold higher risk of CKD compared to population average (OR 5.17, 95%CI: 2.94-9.08, P=1.0E-08). In summary, we demonstrate that the penetrance of kidney disease in T62P carriers is strongly modified by both age and polygenic risk.

## Introduction

Chronic kidney disease (CKD) represents a global health problem, affecting 10-15% of the population and contributing to morbidity and mortality^1^. CKD is highly heritable and has a complex genetic determination with both rare and common allelic contributions^2–4^. Monogenic disorders account for up to 9.3% of all-cause CKD, with autosomal dominant polycystic kidney disease (ADPKD), collagen 4-alpha-associated nephropathies (COL4A-AN) accounting for the most common inherited forms^5,6^. These disorders are characterized by incomplete penetrance and variable progression to kidney failure, including inconsistent intra-familial expressivity among individuals carrying the same pathogenic variant^7^.

Autosomal dominant tubulointerstitial kidney disease (ADTKD) represents the third most common form of monogenic kidney disease^5,6^. The majority of ADTKD cases are caused by pathogenic missense variants in the *UMOD* gene. In contrast to ADPKD and COL4A-AN, these variants are ultra-rare and highly penetrant, leading to uromodulin misfolding and trapping in the endoplasmic reticulum, and causing severe tubular toxicity under dominant inheritance^8^. More recently, a novel intermediate-effect size *UMOD* variant, p.Thr62Pro (T62P, rs143248111), has been described in association with ADTKD^9^. This variant is present in approximately 1 in 1,000 individuals of European ancestry; however, due to incomplete disease penetrance, only a fraction of carriers will express features of CKD. Compared with canonical ADTKD mutations, those T62P carriers that develop CKD displayed reduced disease severity, with slower progression of CKD and an intermediate reduction of urinary uromodulin levels, in line with the milder mutant uromodulin trafficking defect demonstrated *in vitro*^9^.

Genome-wide association studies (GWAS) have discovered hundreds of common variants across the human genome associated with kidney function^10,11^. These discoveries empowered development of genome-wide polygenic risk scores (GPS) for kidney disease, which estimate inherited risk based on aggregate common genetic variation^12,13^. Emerging evidence suggests that polygenic risk can modulate the penetrance and expressivity of monogenic disorders such as familial hypercholesterolemia, hereditary breast and ovarian cancer, and Lynch syndrome^14^. We have recently demonstrated that polygenic score also affects the risk of kidney disease in ADPKD and COL4A-AN^15^. Among the top GWAS loci for kidney function, there were small-effect high-frequency variants at the *UMOD* locus associated with increased *UMOD* expression and higher risk of CKD^10^. An interaction of these regulatory variants with age has also been described^16,17^.

In this study, we aimed to determine whether age or polygenic risk contributes to the highly variable penetrance of the *UMOD* T62P variant. For this purpose, we leveraged exome/genome sequencing and electronic health record (EHR) data for 882,306 individuals across three major biobanks (see study workflow in **Suppl. Figure 1**). In the discovery stage, we analyzed 469,835 participants of the UKBB to test if polygenic risk alters the risk of CKD among T62P carriers. We validated our results in 412,508 participants of the All of Us Research Program and the Geisinger MyCode Community Health Initiative. We generated stably transfected kidney tubular cells expressing wild-type (WT) vs. T62P and canonical ADTKD *UMOD* variants to analyze the processing and maturation of uromodulin and the impact of aging on cellular stress and tubular damage. Our data support age-dependent penetrance related to tubular cell toxicity of the *UMOD* T62P variant due to a protein processing defect, and a significant interaction of T62P with polygenic risk. These findings highlight an interplay between intermediate effect variants, common variant polygenic effects, and aging in the risk of CKD.

## Methods

### Ethics statement

This study analyzes fully de-identified biobank data with all participants consented for genetic studies. Our protocol was reviewed and approved by the Columbia University Institutional Review Board (Protocol AAAC7385).

### Study Design

This cross-sectional study was conducted in two stages. In the discovery stage, we analyzed participants from the UK Biobank (UKBB, N = 469,835), the largest uniform genetic dataset with both SNP array and exome sequencing data available. In the replication stage, we combined electronic health records data from the All of Us Research Program (AoU, N = 245,388) and the Geisinger Community Health Initiative (MyCode, N = 167,120)^18,19^.

### T62P Variant

The primary variant of interest was the c.184A>C, p.Thr62Pro (T62P) missense variant in *UMOD* (rs143248111-G). Carrier status was defined as having at least one copy of the risk allele (i.e., dominant coding). Variant calls were extracted from exome or genome sequence datasets; the variant passed basic quality control assessments in each biobank, including genotype quality (GQ) > 90 and depth of coverage (DP) > 10. The allelic frequencies across all ancestries were 0.143% in UKBB, 0.085% in AoU, and 0.044% in MyCode biobanks.

### Genome-wide Polygenic Score (GPS)

To test the effects of polygenic risk, we used our previously validated genome-wide polygenic score for CKD^12^. The GPS was computed from imputed genotypes using PLINK (--score function), standard normalized, and ancestry-adjusted using the 1000 Genomes reference panel as previously described^12,20^. Because the original optimization of this GPS involved a portion of the UKBB participants, including individuals with a *UMOD* T62P variant, for sensitivity analyses, we excluded all T62P carriers from the optimization step and re-derived the GPS using the same procedure as described in our initial report^12^. The optimal model remained unchanged, confirming that our risk estimates among T62P carriers were not biased by GPS training (**Suppl. Table 1**). To test if common regulatory variants at the *UMOD* locus drive any of the effects of GPS among T62P carriers, we additionally recalculated the GPS after excluding variants within ±200 kb of the *UMOD* gene (**Suppl. Table 2**).

### CKD phenotyping and case-control definitions

We defined CKD cases using our validated e-phenotyping algorithm^2,21^. CKD cases were defined by eGFR <60 ml/min/1.73 m² by the 2021 CKD-EPI equation (CKD stage 3 or above) or kidney failure defined by the need for renal replacement therapy. Controls were defined by eGFR >90 ml/min/1.73 m² and no evidence of CKD based on EHR diagnostic and procedure codes. Similar to our prior studies, we excluded individuals with eGFR 60-90 ml/min/1.73 m² to reduce any potential age-related misclassification of CKD case status^3^.

### Generation of stable cell lines, cell culture and treatments

A pcDNA3.1 expression vector encoding wild-type (WT) human *UMOD* with an N-terminal Strep-tag under a CMV promoter was kindly provided by Gregor Weiss (ETH, Zurich). Single point mutations were introduced by site-directed mutagenesis using the QuickChange Lightning mutagenesis kit (Agilent Technologies, 210515) to obtain the human *UMOD* variants coding for T62P and R185S mutant uromodulin using the mutagenic primers 5′-GGCGATGGCCTGCCCTGCGTGGACC-3′ and 5′-TGGACGAGTACTGGAGCAGCACCGAGTAC-3′, respectively (Microsynth AG). Plasmids were linearized using the restriction enzyme *Bgl*II (New England Biolabs). Mouse inner medullary collecting duct (mIMCD3, ATCC CRL-2123) cells that lack endogenous uromodulin were used to generate stable cell lines expressing *UMOD* variants. mIMCD3 cells were transfected with the linearized constructs using Lipofectamine 3000 (Thermo Fisher Scientific) according to the manufacturer’s instructions to facilitate genomic integration via non-homologous recombination. 48 hours after transfection, cells were subjected to selection with 0.45 mg/mL G418 antibiotics (Sigma Aldrich) for 10 days. Surviving colonies were isolated and single cells were seeded into 96-well plates (Costar, 3595), expanded to 24-well plates (Sarstedt, 83.3922), and screened for uromodulin expression by Western blotting.

Clones exhibiting stable uromodulin expression were cultured in Iscove’s modified Dulbecco’s medium (IMDM, Thermo Fisher Scientific, Waltham, MA, USA) supplemented with 10% fetal bovine serum (FBS, Thermo Fisher Scientific) and 1% Penicillin/Streptavidin (Thermo Fisher Scientific), at 37 °C and 5% CO₂. To preserve selection pressure, cultures were passaged in medium containing 0.1 mg/mL G418. The genomic integration of the full *UMOD* transcript was confirmed by PCR on genomic DNA isolated from each clone and subsequent sequencing. Similar *UMOD* transcript levels in WT and T62P expressing cells were confirmed (mean c_t_-value: *UMOD*-WT, 25.08; *UMOD*-T62P, 24.88). Clonal stability was monitored for up to 30 passages by repeated assessment of uromodulin expression.

To determine tolerable treatment conditions, cells were treated with increasing concentrations of dithiothreitol (DTT) up to 20 mM, and cell viability following DTT exposure was assessed using a 3-(4,5-Dimethylthiazol-2-yl)-2,5-Diphenyltetrazolium Bromide (MTT) assay. Time-course experiments were subsequently performed by exposing cells to 1 mM of DTT for 2, 4, 6, or 8 hours. To induce ER-stress, cells were incubated with 1 mM dithiothreitol (DTT) for 8 h. RNA and protein samples were collected and analyzed by Western blot, RT-qPCR, and immunofluorescence. To investigate the effect of aging, cells were harvested at increasing passage numbers before Western blot and RNA sequencing.

Telomere length was assessed at different cell passages using the quantitative PCR method as previously described^22^. Primer sequences are listed in **Suppl. Table 3**.

### Protein sample preparation and immunoblotting

Cells were lysed in radioimmunoprecipitation assay buffer (RIPA, Sigma-Aldrich) supplemented with protease and phosphatase inhibitors (Sigma-Aldrich) and detached using cell scraper (99002, TPP Techno Plastic Products, Trasadingen, Switzerland). The lysates were centrifuged for 15 min at 1000 g and 4 °C to remove debris. Protein concentrations were determined using the bicinchoninic acid (BCA) protein assay kit (Thermo Fischer Scientific). For glycosidase assays, 20 µg of protein were denatured at 95 °C for 10 min and digested with Endo H (New England Biolabs) or PNGase F (New England Biolabs) for 1 h at 37 °C according to manufacturer instructions and subsequently analyzed by Western blotting. For immunoblotting, protein lysates were reduced with DTT (except for uromodulin) and denatured by boiling at 95°C for 5min. Proteins were separated on 7.5% 12% SDS–PAGE gels under non-reducing conditions for UMOD and under reducing conditions for all other targets. Proteins were transferred to methanol-activated PVDF membranes (Biorad, 1620177) using either wet transfer (UMOD) or semi dry TurboTransfer (other proteins). Membranes were blocked in 5% skimmed milk or 3% BSA (phospho-PERK) for 1 h at room temperature and incubated with appropriate primary antibodies overnight at 4 °C: sheep anti-uromodulin (1:500), rabbit anti-GRP78/BiP (1:1000), rabbit anti-Hsc70 (1:1000), goat anti-LCN2 (1:500), rabbit anti-p21 (1:1000), rabbit anti-p16 (1:1000), rabbit anti-eIF2α (1:500), rabbit anti-p-eIF2α (1:500), rabbit anti-PERK (1:500), rabbit anti-p-PERK (1:500), rabbit anti-ATF4 (1:500), and mouse anti–β-actin (1:10 000). After washing in PBS□T, membranes were incubated with species-specific HRP-conjugated secondary antibodies (1:5000) for 1 h at room temperature. Blots were developed using Bio-Rad ECL substrates (Max ECL for UMOD and LCN2; standard ECL for all other proteins) and imaged on a ChemiDoc system. Immunoblots were quantified by densitometric analysis using ImageJ (version 1.54p, Java 1.8.0_172). Protein quantifications were normalized over β-actin. A list of primary antibodies used in this study is available in **Suppl. Table 4**.

### RNA isolation, reverse transcription, and quantitative PCR

Total RNA was extracted from cells using Direct-zol RNA MicroPrep Kit (Zymo Research) following the manufacturer’s protocol. Reverse transcriptase reaction was executed with iScript TM cDNA Synthesis Kit (Bio-Rad) with up to 1 µg of RNA. The variations in mRNA levels of the target genes were established by relative RT-qPCR with a CFX96TM Real-Time PCR Detection System and the iQ™ SYBR Green Supermix (Bio-Rad) for the detection of single PCR product accumulation. Target-specific primers were designed with Primer3 (web version 4.1.0; Institute of Computer Science, University of Tartu, EST). The PCR conditions were: 95 °C for 3 min, followed by 40 cycles of 15 seconds at 95 °C and 30 seconds at 60 °C. Gene expression was normalized over *Gapdh*. The relative changes were determined by the formula: 2^-ΔΔ*Ct*^ and expressed as relative to UMOD-WT or untreated cells. Primer sequences used can be found in **Suppl. Table 3**.

### RNA-sequencing and bioinformatic analysis

RNA sequencing of cell lysates from *UMOD*-T62P expressing cells was performed at the Functional Genomics Center Zurich (ETH and UZH, Zurich, Switzerland) using an Illumina NovaSeq X Plus sequencer (Illumina, San Diego, CA, USA). The RNA-seq data processing and analysis were performed on SUSHI (Hatakeyama et al., 2016) using the DESeq2 algorithm (Love et al, 2014). Generation of heat maps and overrepresentation analysis (ORA) was done on ExploreDE^23^. Volcano plots and ORA scatterplots were generated with GraphPad Prism.

### Immunofluorescence and image analysis

Cells were seeded in single-chamber dishes (Vitaris/Ibidi, 81156-400-IBI) and fixed either with cold methanol (–20 °C, 10 min) or, for calnexin staining, with 4% paraformaldehyde (PFA, 15 min, RT). Samples were permeabilized and blocked in 0.5% saponin/0.5% BSA for 1 hour. Primary antibodies were applied overnight at 4°C: sheep anti-uromodulin (1:600), rabbit anti-calnexin (1:400), and rabbit anti-GRP78/BiP (1:400). Corresponding Alexa Fluor–conjugated secondary antibodies (1:300) and DAPI (1:1000) were added for 1 h at RT. Samples were mounted with ProLong Gold Antifade Reagent (Thermo Fisher Scientific, P36931) and analyzed utilizing a Leica SP8 confocal laser scanning microscope (Leica Microsystems GmbH, Wetzlar, Germany) with a 63× oil objective (NA 1.4). Sequential scanning was used to avoid spectral overlaps. Quantitative image analysis was performed by randomly selecting 10 visual fields, each containing 10-15 cells, pooled from 3 biological replicates. Images were analyzed in ImageJ/Fiji, and colocalization was quantified using the Colocalization plugin.

### Statistical Analysis

In all experiments, measured values were expressed as the mean ± SEM. Before statistical analysis, outliers were identified using the robust regression and outlier removal (ROUT) method and excluded from subsequent analyses. Residuals were evaluated for normality and homogeneity of variances (F test). Two-group comparisons were performed using an unpaired, two-tailed Student’s t-test. Comparisons between three or more groups were performed using ordinary one-way ANOVA followed by Tukey-Kramer’s post hoc analysis or Šídák’s multiple comparisons test. When variances were unequal, Welch’s one-way ANOVA followed by Dunnett’s T3 multiple comparisons test was applied. P < 0.05 was considered as statistically significant. The specific statistical tests applied to each dataset are indicated in the corresponding figure legends; statistical significance with thresholds defined as *P < 0.05, **P < 0.01, ***P < 0.001, and ****P < 0.0001. GraphPad Prism v.10 was used to visualize the data.

To test genetic predictors, such as T62P or GPS, against the outcome of CKD (case/control) in biobank datasets, we used logistic regression. All models were adjusted for age, sex, diabetes, genotyping batch (if applicable), and the first 5 principal components of genetic ancestry. To test GPS effects among carriers, we performed analyses stratified by T62P carrier status. The association between the GPS (continuous predictor) and CKD was expressed as an odds ratio (OR) per one standard deviation (SD) of the GPS distribution. To examine the effect of age on our effect estimates, we performed sensitivity analyses using varying age inclusion thresholds (over 40, 45, 50, 55, 60, and 65 years). Models were fitted separately within each age-defined subgroup using the same methods as above. To test for multiplicative interaction between GPS and T62P carrier status, we included an interaction term in the logistic regression model along with the main predictors. To estimate CKD risk by quintiles of GPS for carriers vs. non-carriers, we used the middle quintile of non-carriers for reference. Logistic regression was then used to estimate ORs for each quintile compared to the reference group. These analyses were implemented in R version 4.3. Meta-analyses across cohorts were conducted using inverse variance-weighted fixed-effects models (METAL 2011-03-25)^24^.

### Meta-phenome wide association studies (Meta-PheWAS)

We conducted a meta-phenome-wide association study (Meta-PheWAS) to test the effects of *UMOD* T62P across all three biobanks. The AoU dataset included 180,712 participants with genome sequence data and 13,334 ICD-9 codes mapped to 1,817 distinct phecodes. The UKBB dataset included 422,695 participants with exome sequence data and 5,613 ICD-9 codes mapped to 1,817 phecodes. The MyCode dataset included 167,120 participants with 45,364 ICD-9 and ICD-10 codes mapped to 1,817 PheCodes. Phenome-wide associations were performed using the PheWAS R package, which defines cases as individuals with at least two occurrences of ICD codes within a given phecode and pre-defined controls for each phecode^25^. Logistic regression was used for each phecode, with case-control status as the outcome and genotype, sex, age, batch, and five principal components of ancestry as covariates. Fixed-effects meta-analysis combining AoU and UKBB results was then performed using the same PheWAS R package. A Bonferroni-corrected significance threshold of P = 2.75 × 10⁻□ (0.05 / 1,817 phecodes) was used to define phenome-wide significance.

## Results

### The T62P *UMOD* variant conveys CKD risk with age-dependent penetrance

We first analyzed exome sequence data from the participants of the UKBB (discovery cohort) to define 638 T62P carriers and 469,160 non-carriers. After electronic phenotyping for CKD using our validated algorithm, we tested the effect of the T62P variant on the risk of kidney disease in this cohort. We confirmed that the T62P variant was associated with a two-fold increased risk of CKD stage 3 or above (OR 2.08, 95%CI: 1.37-3.13, P=4.9E-04) and a 3-fold increased risk of kidney failure (OR 3.22, 95%CI: 1.19-8.72, P=2.1E-02) under a dominant inheritance model (**Table 1**).

**Table 1.** The effects of T62P and GPS on the risk of (a) CKD and (b) kidney failure in the discovery, replication, and all cohorts combined. The effects estimated for each predictor (T62P variant, GPS among T62P carriers, and GPS among T62P non-carriers) are adjusted for age, sex, diabetes, array batch (if applicable), and genetic ancestry. The discovery cohort was based on the UK Biobank (638 carriers and 469,160 non-carriers). The replication cohort combined data from the All-of-Us Study and the MyCode biobank (AoU=209 carriers and AoU=245,179 non-carriers; MyCode=155 carriers and 166,965 non-carriers). The combined analysis includes meta-analysis of all three biobanks (UK Biobank, All-of-Us, and MyCode: 1,002 carriers and 881,304 non-carriers).

**A. Risk of CKD**
|  | **T62P Variant**<br>**OR (95%CI), P-value** | **GPS in T62P Carriers**<br>**OR per SD (95%CI), P-value** | **GPS in Non-Carriers**<br>**OR per SD (95%CI), P-value** |
| --- | --- | --- | --- |
| Discovery | 2.08 (1.37-3.13), P=4.9E-04 | 4.68 (3.03-7.11), P=8.9E-05 | 1.83 (1.81-1.85), P<E-300 |
| Replication | 1.06 (0.68-1.68), P=7.8E-01 | 2.57 (1.21-5.47), P=1.4E-02 | 1.44 (1.42-1.46), P<E-300 |
| Combined | 1.54 (1.13-2.09), P=5.6E-03 | 3.44 (2.50-4.73), P=7.1E-06 | 1.56 (1.55-1.57), P<E-300 |

**B. Risk of Kidney Failure**
|  | **T62P Variant**<br>**OR (95%CI), P-value** | **GPS in T62P Carriers**<br>**OR per SD (95%CI), P-value** | **GPS in Non-Carriers**<br>**OR per SD (95%CI), P-value** |
| --- | --- | --- | --- |
| Discovery | 3.22 (1.19-8.72), P=2.1E-02 | 5.47 (0.98-30.5), P=5.2E-02 | 1.26 (1.19-1.34), P=7.1E-14 |
| Replication | 2.62 (1.27-5.42), P=8.5E-03 | 2.03 (0.73-5.63), P=1.7E-01 | 1.25 (1.19-1.31), P=1.2E-14 |
| Combined | 2.79 (1.57-4.98), P=4.9E-04 | 2.59 (1.07-6.23), P=3.4E-02 | 1.25 (1.20-1.30), P=1.2E-32 |

To assess the effect of age on the penetrance of the T62P variant in UKBB, we performed the T62P association analysis for a range of age inclusion thresholds (**Table 2**). We observed a consistent trend for increased T62P risk effect with older age. For example, the T62P variant was associated with over 3.3-fold increased risk of CKD among individuals over 65 years of age (OR 3.35, 95%CI: 1.82-6.17, P=1.0E-04) as compared to a 2-fold increased risk for those over 40 years of age (OR 2.08, 95%CI: 1.37-3.13, P=4.9E-04).

**Table 2.** Age-dependent effects of T62P and GPS on the risk of (a) CKD and (b) kidney failure in the UKBB. The effects were estimated for each predictor (T62P variant, GPS among T62P carriers, and GPS among T62P non-carriers) at variable age inclusion thresholds. All models were adjusted for age, sex, diabetes, array batch, and genetic ancestry. GPS effect estimates for carriers in the age group >65 years are missing due to small case counts in this group.

**A. Risk of CKD**
| Age (years) | T62P Variant<br>OR (95%CI), P | GPS in T62P Carriers<br>OR per SD (95%CI), P | GPS in Non-Carriers<br>OR per SD (95%CI), P |
| --- | --- | --- | --- |
| >40 | 2.08 (1.37-3.13), P=4.9E-04 | 4.81 (2.20-10.5), P=8.1E-05 | 1.85 (1.81-1.89), P<E-300 |
| >45 | 2.11 (1.40-3.19), P=3.7E-04 | 4.81 (2.20-10.5), P=8.1E-05 | 1.86 (1.81-1.90), P<E-300 |
| >50 | 2.20 (1.45-3.34), P=1.9E-04 | 4.81 (2.20-10.5), P=8.1E-05 | 1.89 (1.84-1.93), P<E-300 |
| >55 | 2.41 (1.58-3.68), P=4.1E-05 | 4.90 (2.23-10.8), P=7.5E-05 | 1.90 (1.85-1.94), P<E-300 |
| >60 | 2.61 (1.67-4.09), P=2.4E-05 | 4.57 (2.02-10.4), P=2.6E-04 | 1.93 (1.88-1.98), P<E-300 |
| >65 | 3.35 (1.82-6.17), P=1.0E-04 | 5.26 (1.56-17.7), P=7.5E-03 | 1.99 (1.92-2.06), P<E-300 |

**B. Risk of Kidney Failure**
| Age (years) | T62P Variant<br>OR (95%CI), P | GPS in T62P Carriers<br>OR per SD (95%CI), P | GPS in Non-Carriers<br>OR per SD (95%CI), P |
| --- | --- | --- | --- |
| >40 | 3.22 (1.19-8.72), P=2.1E-02 | 5.47 (0.98-30.5), P=5.2E-02 | 1.26 (1.19-1.34), P=7.1E-14 |
| >45 | 3.42 (1.26-9.28), P=1.5E-02 | 5.47 (0.98-30.5), P=5.2E-02 | 1.27 (1.19-1.35), P=3.8E-14 |
| >50 | 3.82 (1.41-10.4), P=8.5E-03 | 7.24 (1.03-51.1), P=4.7E-02 | 1.26 (1.18-1.35), P=3.1E-12 |
| >55 | 4.76 (1.74-13.0), P=2.4E-03 | 7.69 (1.00-59.1), P=4.8E-02 | 1.26 (1.17-1.35), P=2.3E-10 |
| >60 | 4.62 (1.44-14.9), P=1.0E-02 | 9.39 (0.27-320), P=2.1E-01 | 1.31 (1.20-1.42), P=5.1E-10 |
| >65 | 8.00 (1.83-35.0), P=5.7E-03 | --- | 1.37 (1.20-1.55), P=1.2E-06 |

### The T62P variant causes impaired uromodulin processing and increased retention in endoplasmic reticulum

To substantiate potential nephrotoxic effects of the T62P variant, we generated stable mouse inner medullary collecting duct (mIMCD3) cell lines expressing similar levels of Strep-tagged wild-type (WT) or mutant (T62P and R185S) uromodulin (**Figure 1a**). Compared to WT cells, clones of T62P-expressing cells consistently exhibited a defective uromodulin maturation profile, evidenced by the presence of a premature precursor band (85 kDa), as well as elevated levels of endoplasmic reticulum (ER) stress marker GRP78 and Hsc70 (**Figure 1b**). Glycosylation analysis revealed that WT, mature uromodulin is Endo H resistant and fully glycosylated, consistent with proper Golgi processing. Conversely, T62P uromodulin was partially sensitive to Endo H, consistent with ER retention, which was less pronounced than that observed for the severe disease-associated R185S uromodulin (**Figure 1c**). Confocal microscopy analysis showed that in WT cells, uromodulin displays a limited overlap with the ER chaperone calnexin (CNX), whereas in T62P-expressing cells, the colocalization of uromodulin with CNX was significantly increased, indicating retention within the ER, less pronounced than the canonical R185S ADTKD mutant (**Figure 1d**).

**Figure 1.**
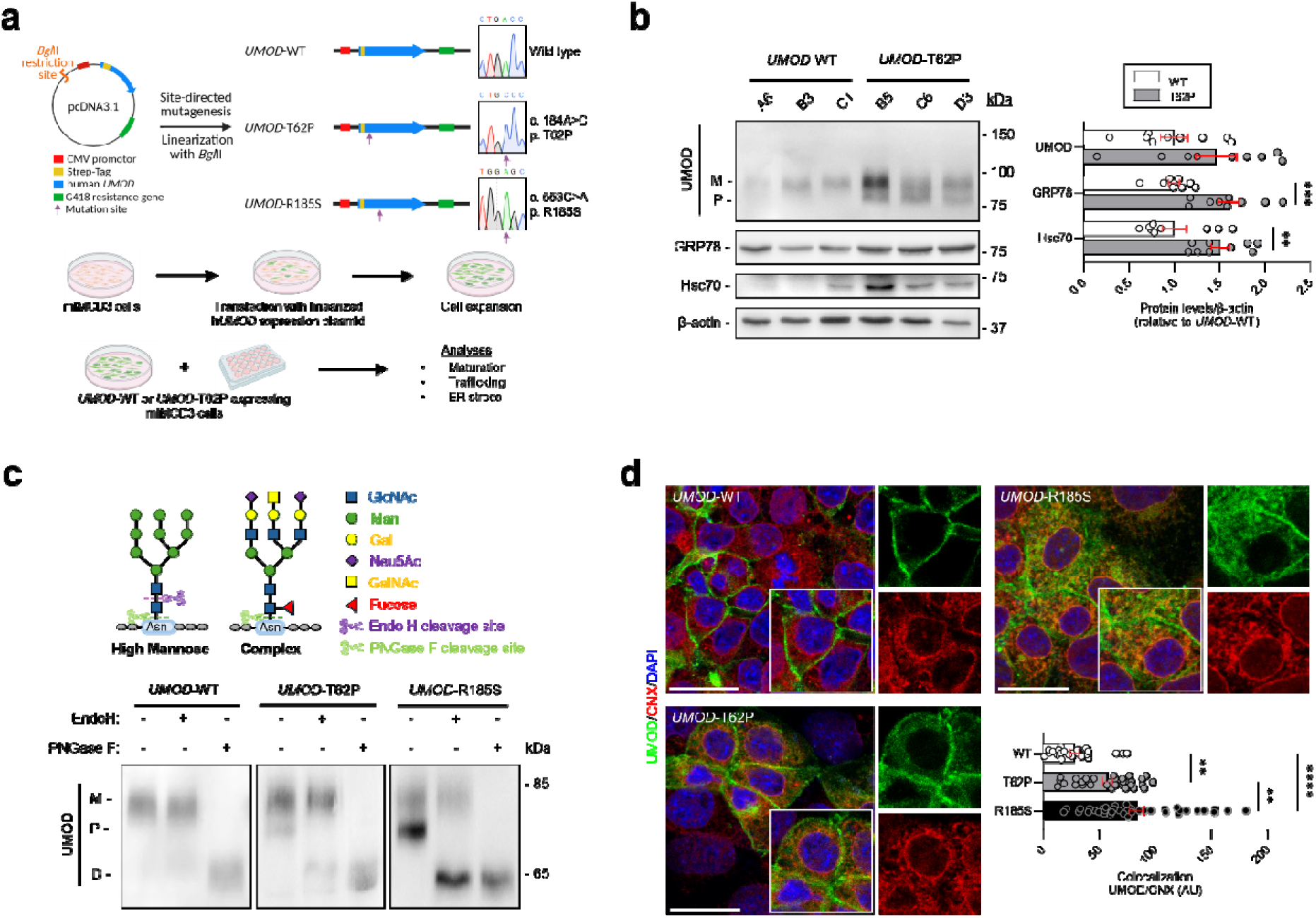
Defective maturation and trafficking of T62P *UMOD* variant in kidney tubular cells. (**a**) Schematic overview of the expression vector and linearized products with indicated point mutations (top). Generation of stable mIMCD3 cell lines expressing wild-type (WT) and T62P and R185S *UMOD* variants and the experimental workflow used for downstream analysis (bottom). (**b**) Representative immunoblots of selected stable clones expressing WT or T62P *UMOD*. Expression of uromodulin (UMOD), GRP78, and Hsc70 is shown. β-actin served as a loading control (n = 9 biological replicates). M: mature; P: precursor. Bars represent mean ± SEM. Unpaired two tailed t test. *P<0.05, **P<0.01, ***P<0.001, ****P<0.0001. (**c**) Top: Schematic of different types of N-linked Glycans with specific cleavage sites of Endo H and PNGase F. Bottom: Representative immunoblots for UMOD of lysates from cells stably expressing WT, T62P, or the severe canonical R185S *UMOD* variants following Endo H or PNGase F digestion. M: mature; P: precursor; D: deglycosylated. (**d**) Immunofluorescence analysis of stable clones expressing WT, T62P and R185S *UMOD* variants, stained for UMOD (green) and calnexin (CXN, red). Nuclei are stained with DAPI. Scale bars, 20 µm. (n>25 cells per condition). Ordinary one-way ANOVA followed by Tukey’s post hoc test. *P<0.05, **P<0.01, ***P<0.001, ****P<0.0001.

### The T62P variant engages increased ER stress and additional cell quality control

The processing of uromodulin in the ER involves the formation of 24 intramolecular disulfide bonds^26^. To assess cellular responses to acute ER stress, WT and T62P cells were treated with different concentrations of dithiothreitol (DTT), a reducing agent that disrupts disulfide bond formation - a critical step in uromodulin folding and maturation (**Suppl. Figure 2**). Exposure to increasing concentrations of DTT (up to 20 mM, 8 hours) did not affect the viability of WT cells, whereas treatment above 2mM significantly impacted T62P cell survival (**Suppl. Figure 2a**). Time-course analysis after exposure to non-toxic 1mM DTT revealed detectable levels of mature uromodulin in WT cells up to 6 hours of treatment, with the appearance of immature UMOD observed only after 8 hours of exposure. T62P-expressing cells however, showed a progressive, time-dependent loss of mature uromodulin coupled to an increase in the immature form, with an almost complete conversion after 8 hours of DTT treatment (**Suppl. Figure 2b**) - indicating enhanced sensitivity and reduced folding resilience of T62P uromodulin, as also marked by increased GRP78 and Hcs70 levels over time.

We next investigated trafficking and processing responses of WT and T62P uromodulin in mIMCD3 cells following treatment with a well-tolerated dose of DTT (**Figure 2a**). Confocal microscopy revealed that treatment with DTT further enhanced the colocalization of T62P uromodulin with GRP78, already higher than WT at baseline, whereas only a small significant change could be detected in WT cells (**Figure 2b**). Immunoblot analysis indicated a reduction from almost exclusive mature uromodulin to about 45% in WT cells upon DTT treatment, whereas T62P cells displayed reduced maturation already at baseline (~20% immature uromodulin), with DTT treatment resulting in near-complete conversion to the immature form (**Figure 2c**). These specific changes were associated with differential induction of ER stress, as indicated by GRP78 and Hsc70 levels (**Figure 2c**). RT-qPCR analysis showed induction of ER stress, involvement of ER folding machinery, and unfolded protein response-associated transcripts following DTT treatment in both WT and T62P cells, with a stronger transcriptional response in T62P-expressing cells (**Figure 2d**). DTT treatment induced robust upregulation of *Hspa5* and *Ddit3* in both genotypes, with significantly stronger induction observed in T62P-expressing cells. Analysis of chaperone-related transcripts revealed significant induction of *Dnajb4* following DTT treatment, whereas *Dnaja4* expression remained unchanged. Notably, *Hsp90aa1* transcription was significantly increased only in DTT-treated T62P cells. These data indicate that DTT induces a coordinated transcriptional ER stress response in both WT and T62P cells. However, T62P cells experience higher ER stress and have a greater ER chaperone demand, consistent with higher misfolded protein load. The induction of the cytosolic chaperone *Hsp90aa1* only in T62P-expressing cells suggests that the stress response in this variant goes beyond the ER and engages additional cell quality control mechanisms.

**Figure 2.**
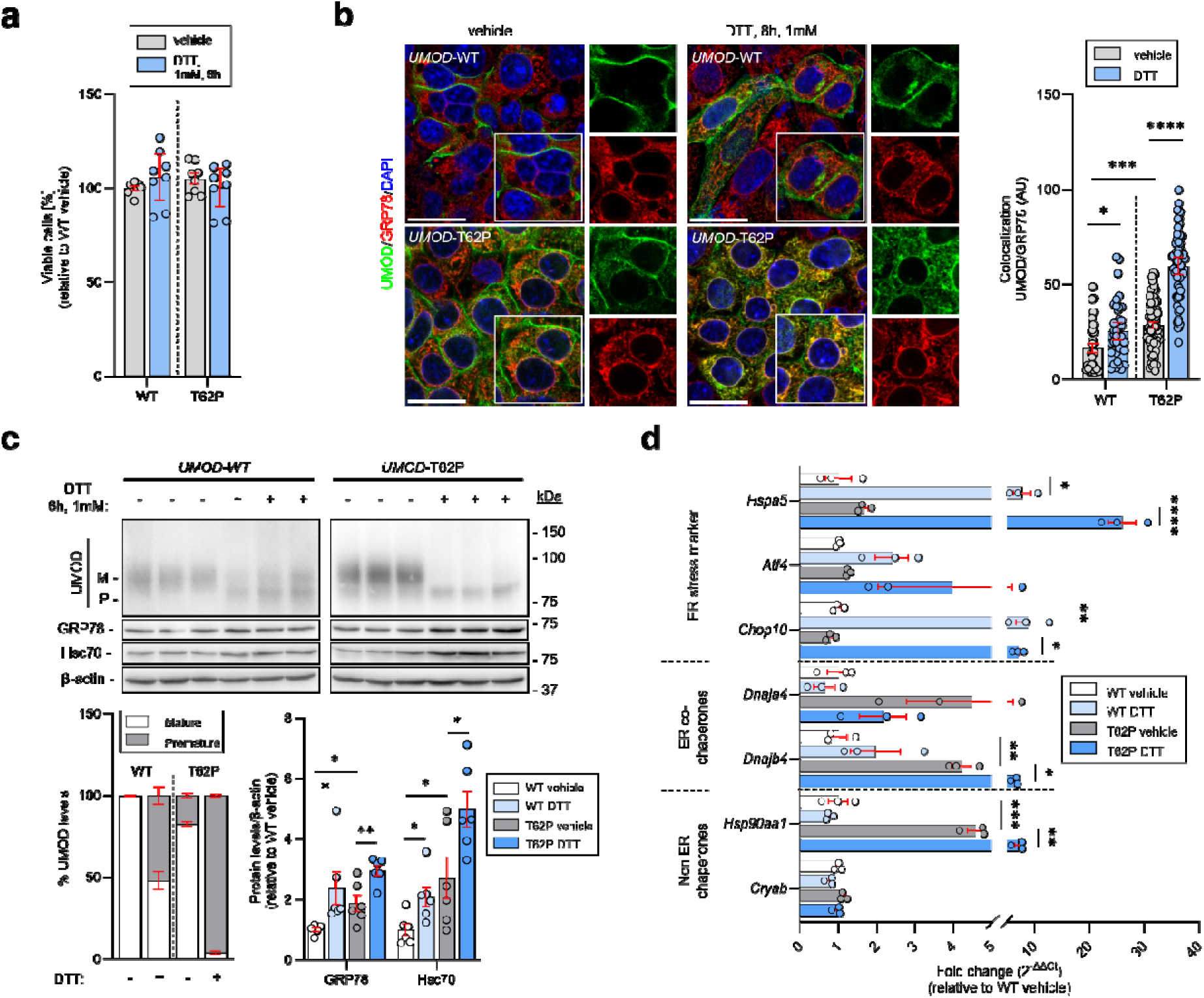
Cellular stress responses in cells expressing wild-type or T62P *UMOD*. (**a**) MTT cell viability of cells stably expressing WT or T62P UMOD under basal conditions (vehicle) or after treatment with 1 mM dithiothreitol (DTT) for 8 h. Bars represent mean ± SEM. Ordinary one-way ANOVA followed by Šídák’s multiple comparisons test. *P<0.05, **P<0.01, ***P<0.001, ****P<0.0001. (**b**) Representative immunofluorescence analysis of *UMOD*-WT or T62P expressing cells for UMOD (green) and GRP78 (red) under basal conditions (vehicle) or after treatment with DTT. Nuclei are stained with DAPI. Scale bar: 20 μm. Welch’s one-way ANOVA followed by Dunnett’s T3 multiple comparisons test (n>31 cells per condition). (**c**) Immunoblot analysis of UMOD, GRP78 and Hsc70. β-actin served as a loading control (n=6 biological replicates). Bars represent mean ± SEM. Unpaired two-tailed t test. *P<0.05, **P<0.01, ***P<0.001, ****P<0.0001. (**d**) RT-qPCR analysis of selected ER-stress, ER co-chaperone or non-ER chaperone genes in cells following treatment with or without DTT (n=3 biological replicates). Bars represent mean ± SEM. Ordinary one-way ANOVA followed by Šídák’s multiple comparisons test. *P<0.05, **P<0.01, ***P<0.001, ****P<0.0001.

### Effect of cellular aging on the deleterious effect of T62P uromodulin

To substantiate the age effect on penetrance of T62P, we tested whether increasing cell passage - a widely used *in vitro* model for cellular aging validated by progressive telomere shortening^27,28^ – impacted the stress responses observed in T62P mIMCD cells (**Figure 3a**; **Suppl. Figure 3a**). RNA-seq profiling at passages 5, 15, and 21 revealed clear passage-dependent transcriptional changes (**Suppl. Figure 3b**). PC1 and PC2 accounted for 68.9% and 20.3% of the total variance, respectively, indicating that passage number is the major source of transcriptional variation. Pairwise transcriptomic comparison of P21 versus P5 (adjusted *p* < 0.05; log_2_FC > 0.5) revealed progressive divergence across passages, and out of the 14,240 transcripts detected, 2,699 (19.0%) were significantly up- and 2,479 (17.4%) significantly down-regulated relative to P5 (**Figure 3b**).

**Figure 3.**
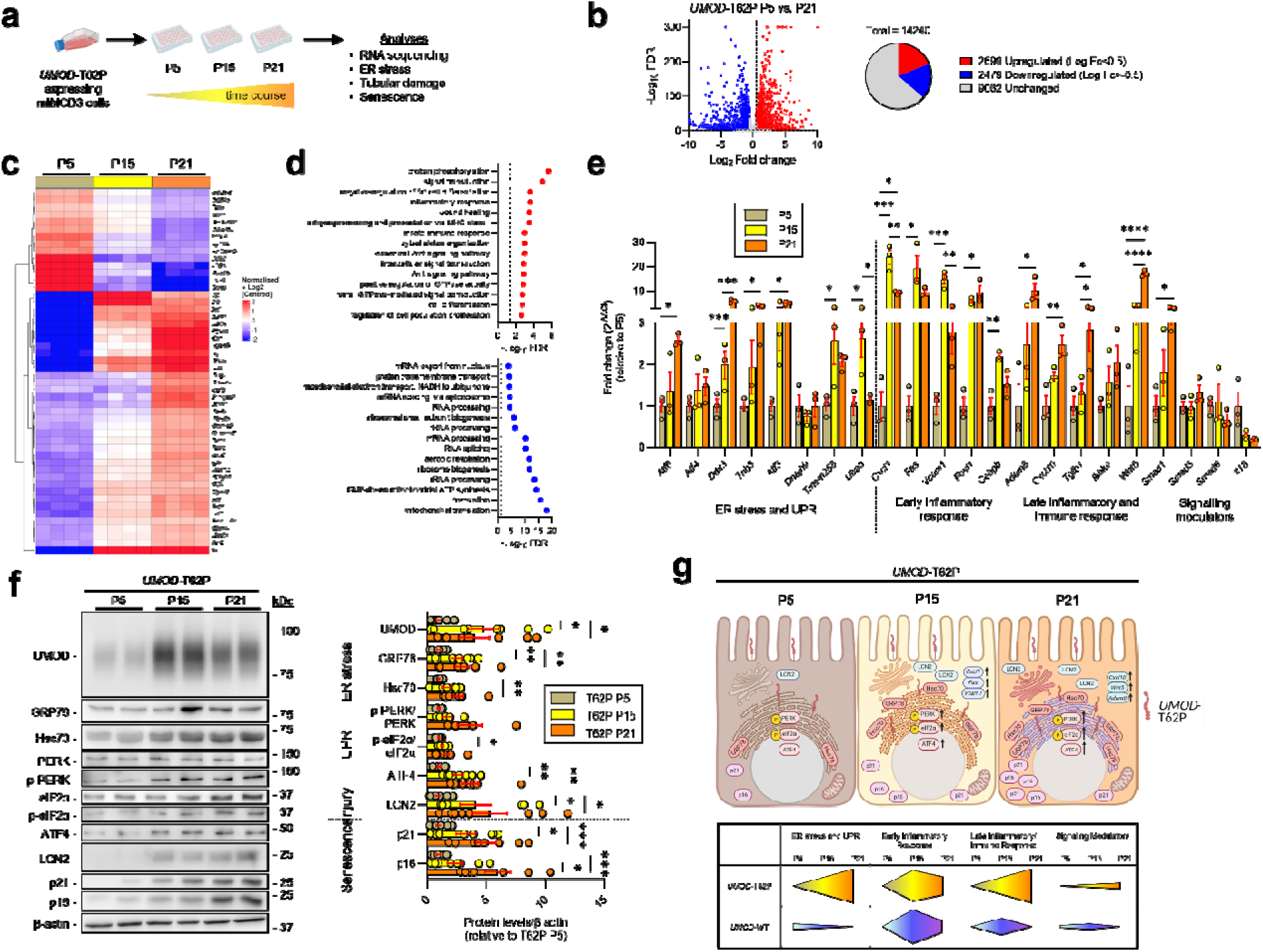
Progressive ER stress, inflammatory remodeling, and senescence-associated adaptation during prolonged culture of T62P-expressing cells. (**a**) Schematic overview of the workflow. (**b**) Volcano plot showing differentially expressed genes (DEGs) between T62P-expressing cells at passages 5 (P5) and 21 (P21). Genes not significantly changed (FDR > 0.05 and Log_2_ FC > 0.5) shown in grey, upregulated genes in red and downregulated genes in blue. Total numbers of unchanged, up-, and downregulated genes are indicated in the pie chart. (**c**) Heatmap of the top 50 DEGs in P21 cells compared to P5 cells. (**d**) Over-representation analysis showing each 15 most significantly up-(red) and downregulated (blue) Gene Ontology (GO) biological processes in T62P P21 versus T62P P5 cells. (**e**) RT-qPCR validation of ER-stress and unfolded protein response (UPR), inflammatory/immune response and signaling modulator genes in *UMOD*-T62P-expressing cells at passages 5, 15 and 21 (n=3 biological replicates). Bars represent mean ± SEM. Ordinary one-way ANOVA followed by Tukey’s post hoc test. *P<0.05, **P<0.01, ***P<0.001, ****P<0.0001. (**f**) Immunoblot analysis of *UMOD*-T62P-expressing cells harvested at increasing passage number (P5, P15, P21), probed for UMOD, GRP78, Hsc70, total and phosphorylated eIF2α and PERK, ATF4, LCN2, p21 and p16. β-actin served as a loading control (n = 5-8 biological replicates). Bars represent mean ± SEM. Unpaired two-tailed t test. *P < 0.05, **P < 0.01, ***P < 0.001, ****P < 0.0001. (**g**) Top: Summary of cumulative effects of cellular aging in T62P-expressing tubular cells across P5, P15 and P21. Bottom: Changes in transcript levels over time in ER stress and UPR; early inflammatory response; late inflammatory and immune response; and signaling modulators, for both *UMOD*-T62P and *UMOD*-WT expressing cells. Arrow-shaped gradients indicate relative magnitude and direction of each response.

Heatmap analysis of selected gene ontology (GO) categories revealed coordinated pathway-level remodeling across passages. Stress-responsive regulators (*Atf3*, *Trib3*, *Ddit3*, *Atf6*) were progressively induced (**Suppl. Figure 3c**), alongside immune and inflammatory genes, including antigen presentation components (*H2-K1*, *H2-D1*, *H2-DMa*, *H2-DMb1*, and *Cd74*) and innate immune regulators (**Suppl. Figure 3d,e**). Inflammatory programs exhibited temporal restructuring: chemokines (*Cxcl1*, *Cxcl5*, *Ccl20*, *Il36a*) peaked at passage 15, whereas later passages showed sustained induction of inflammatory effectors (*Nos2*, *Gsdmd*, *Casp4*, *Pik3cd*, *Adam8*, *Wnt5a*, *Axl*) (**Suppl. Figure 3e**). Highly upregulated genes included key immune and inflammatory regulators (*C3*, *Gbp9*, *Lgals9*, *H2-K1*) and receptors (*Axl*, *Lrp1*, *Igf2r*), reflecting activation of complement, interferon, antigen presentation, and growth factor pathways. Conversely, genes enriched at P5 (*Rab25*, *Cd82*, *Tmc4*) declined with passage, indicating reduced membrane dynamics and extensive structural and functional cellular remodeling (**Figure 3c**). Enrichment analysis confirmed upregulation of immune and inflammatory pathways, along with Wnt signaling and intracellular signal transduction (**Figure 3d**). In contrast, downregulated transcripts were predominantly associated with mitochondrial function and biosynthetic activity, indicating a shift from metabolic and anabolic processes toward inflammatory activation and stress-responsive signaling during cellular aging (**Figure 3d**).

Representative genes underlying these pathway shifts were validated (**Figure 3e**). ER stress-associated genes (*Atf3, Trib3, Ddit3, Atf6*) showed progressive induction across passages in T62P-expressing cells, whereas WT control cells displayed stable or slightly reduced expression levels of these transcripts over time. Together, the distinct profiles indicated selective activation of stress-adaptive pathways in the presence of mutant uromodulin (**Figure 3g**; **Suppl. Figure 3f**). Inflammatory transcripts displayed distinct temporal regulation: *Cxcl1, Fas,* and *Vcam1* peaked at passage 15, whereas *Adam8, Tgfb1, Cxcl10,* and *Wnt5a* remained elevated or further increased, consistent with a shift from early activation toward inflammatory remodeling (**Figure 3e**). Of note, WT cells also exhibited transient induction of selected inflammatory mediators during passaging. However, expression levels largely declined by passage 21, suggesting partial adaptation to replicative stress in the absence of mutant uromodulin. In contrast, inflammatory and stress-associated programs in T62P cells remained persistently activated during prolonged culture (**Figure 3e,g**; **Suppl. Figure 3f**). Western blot analysis (**Figure 3f**) confirmed increased uromodulin accumulation, induction of ER stress markers (GRP78, Hsc70) and components of PERK-associated unfolded protein response (UPR), indicating chronic ER stress and altered proteostasis in T62P cells. These changes were paralleled by increased tubular damage marker LCN2 and senescence markers p21 and p16.

Collectively, these data indicated a mild but significant defect in trafficking and maturation of T62P uromodulin, causing baseline ER stress that is exacerbated under reducing conditions. Cellular aging shifted T62P-expressing cells to a stress-adaptive, inflammatory, and remodeling phenotype, with increased proteostatic burden potentially promoting long-term tubular damage (**Figure 3g**).

### Polygenic risk modifies the penetrance of the T62P variant

To examine if polygenic factors contribute to the variable penetrance of the T62P variant, we next tested the effects of polygenic risk (as captured by GPS) on the risk of CKD stratified by T62P carrier status in the UKBB-based discovery stage. The GPS was a significant predictor of CKD among both T62P carriers (OR per SD 4.68, 95%CI: 3.03-7.11, P=8.9E-05) and non-carriers (OR per SD 1.83, 95%CI: 1.81-1.85, P<E-300), but the risk effect was notably stronger among the carriers (**Table 1**). These effects were independent of age, sex, diabetes, or genetic ancestry.

We also observed a progressive increase in the GPS effect with older age (**Table 2**), and a significant age-by-GPS interaction among the T62P carriers (P=2.1E-08). In T62P carriers that were 65 years or older, the GPS was associated with over a 5-fold higher risk per SD (OR 5.26, 95%CI: 1.56–17.7, P=7.5E-03), suggesting a strong age-dependent amplification of GPS effects.

We also explored if the GPS effect was mediated by known regulatory risk variants at the *UMOD* locus, known to have cis-regulatory effects on *UMOD* gene expression. For this purpose, we reformulated the GPS after excluding all variants within ±200 kb of the *UMOD* coding sequence (**Suppl. Table 2**). The GPS remained strongly associated with CKD risk among carriers after exclusion of the locus and its flanking regions (OR per SD 4.66, 95%CI: 2.15-10.1, P=8.9E-05), suggesting that the observed risk associations are not solely driven by cis-regulatory variants, but rather represent a true genome-wide polygenic effect.

### Replication and meta-analyses across biobanks

To validate these results in an adequately powered independent sample of T62P variant carriers, we combined data from two additional biobanks: AoU and MyCode (**Table 1**). We defined 209 T62P carriers in AoU and 155 in MyCode. In the meta-analysis of both validation cohorts, we independently confirmed that the GPS was a significant predictor of CKD among both carriers (OR per SD 2.57, 95%CI: 1.21 – 5.47, P=1.4E-02) and non-carriers (OR per SD 1.44, 95%CI: 1.42 – 1.46, P<E-300).

We then analyzed data jointly across UKBB, AoU, and MyCode biobanks. First, we tested *UMOD* T62P variant carriers for disease associations phenome-wide using a Meta-PheWAS approach (**Figure 4, <u>Suppl. Data 1</u>**). We confirmed phenome-wide significant association of T62P with the phecode of End-Stage Renal Disease (OR_age ≥40_=4.34, 95%CI: 2.36-7.93, P = 2.1E-06). This association became stronger at a higher age inclusion threshold (OR_age ≥55_ =5.71, 95%CI: 3.10–10.53, P = 2.4E-08). Thus, the Meta-PheWAS validated our results using an alternative phenotyping strategy and confirmed age-dependent penetrance of T62P.

**Figure 4.**
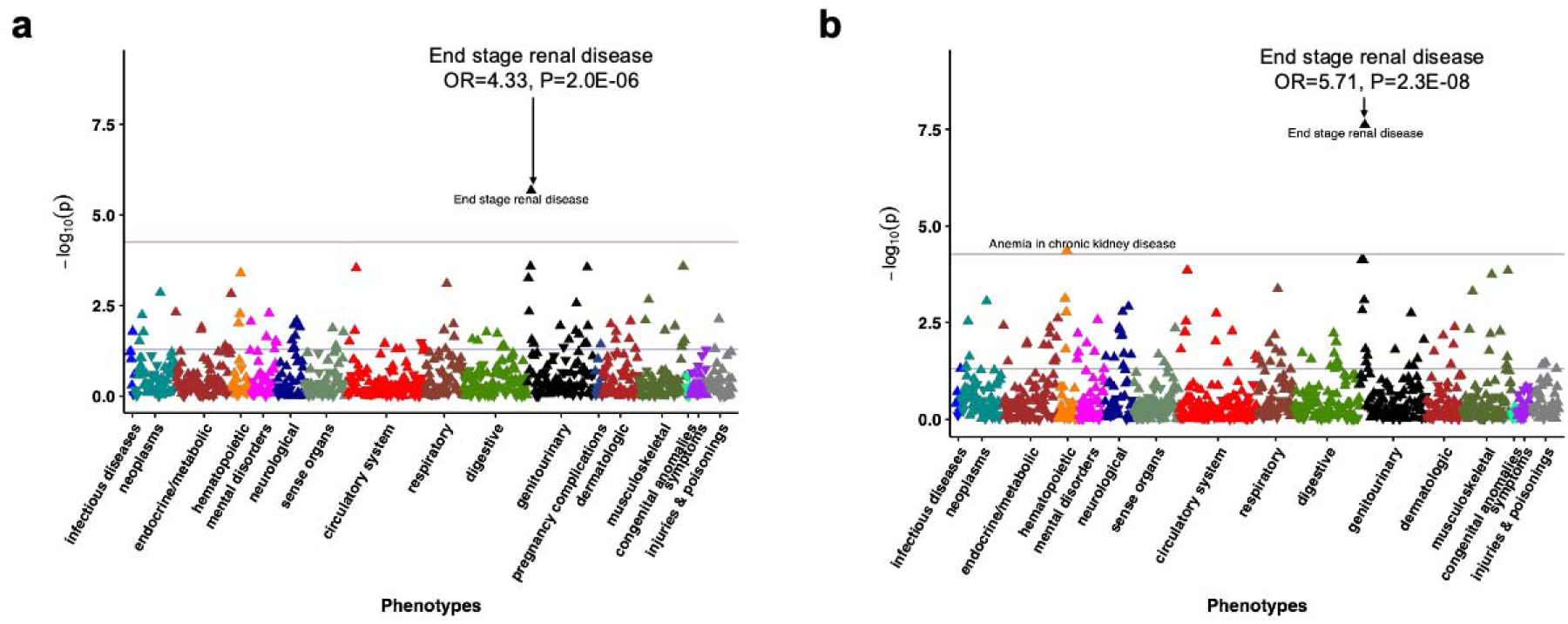
Phenome-wide meta-analysis (Meta-PheWAS) for *UMOD* T62P. Meta-PheWAS of UMOD T62P stratified by age: (a) participants aged ≥40 years and (b) participants aged ≥55 years to examine the effect of age on *UMOD* T62P associations. Analyses include combined data from 422,695 UKBB, 180,712 All of Us, and 167,120 participants with both genotype and phenotype data available. Logistic regression was performed under a dominant inheritance model, adjusted for age, sex, batch, and ancestry. Effect estimates and two-sided P-values were combined across cohorts using fixed-effects meta-analysis. Red horizontal lines indicate the phenome-wide significance threshold accounting for multiple testing (P=2.8×10⁻□). Y-axis: −log_10_(P-value) from fixed-effects meta-analysis (two-sided). X-axis: system-based phecode groupings. Upward-pointing triangles indicate increased odds for a given phecode, and downward-pointing triangles indicate reduced risk.

Next, we assessed the effects of GPS in the pooled analysis of all three biobanks. The GPS increased the risk of CKD by over 3.4-fold per standard deviation among T62P carriers (meta-analysis OR 3.44 per SD, 95%CI: 2.50-4.73, P=7.1E-06, **Table 1**). This effect was more than 2-fold greater compared to non-carriers (meta-analysis OR 1.56 per SD, 95%CI: 1.55-1.57, P<1E-300), suggesting an effect modification. Accordingly, we tested for the GPS-by-T62P interaction in each cohort, followed by meta-analysis of the interaction terms. We observed a significant multiplicative interaction (meta-analysis OR=1.53, 95%CI: 1.02-2.31, P=3.8E-02), confirming GPS-by-T62P effect modification.

To facilitate interpretation of T62P effects in the context of variable polygenic risk of CKD, we expressed T62P effects by GPS quintiles using non-carriers in the middle quintile (reflective of average population risk) as reference, and adjusting for age, sex, diabetes, and genetic ancestry (**Figure 5** and **Suppl. Table 5**). Among the T62P variant carriers, those in the highest GPS quintile had over a 5-fold higher risk of CKD (meta-analysis OR=5.17, 95%CI: 2.94-9.08, P=1.0E-08) compared to average population risk. In contrast, the T62P carriers in the lowest GPS quintile had no statistically higher risk compared to the population average (OR=0.43, 95%CI 0.11-1.73, P=2.4E-01).

**Figure 5.**
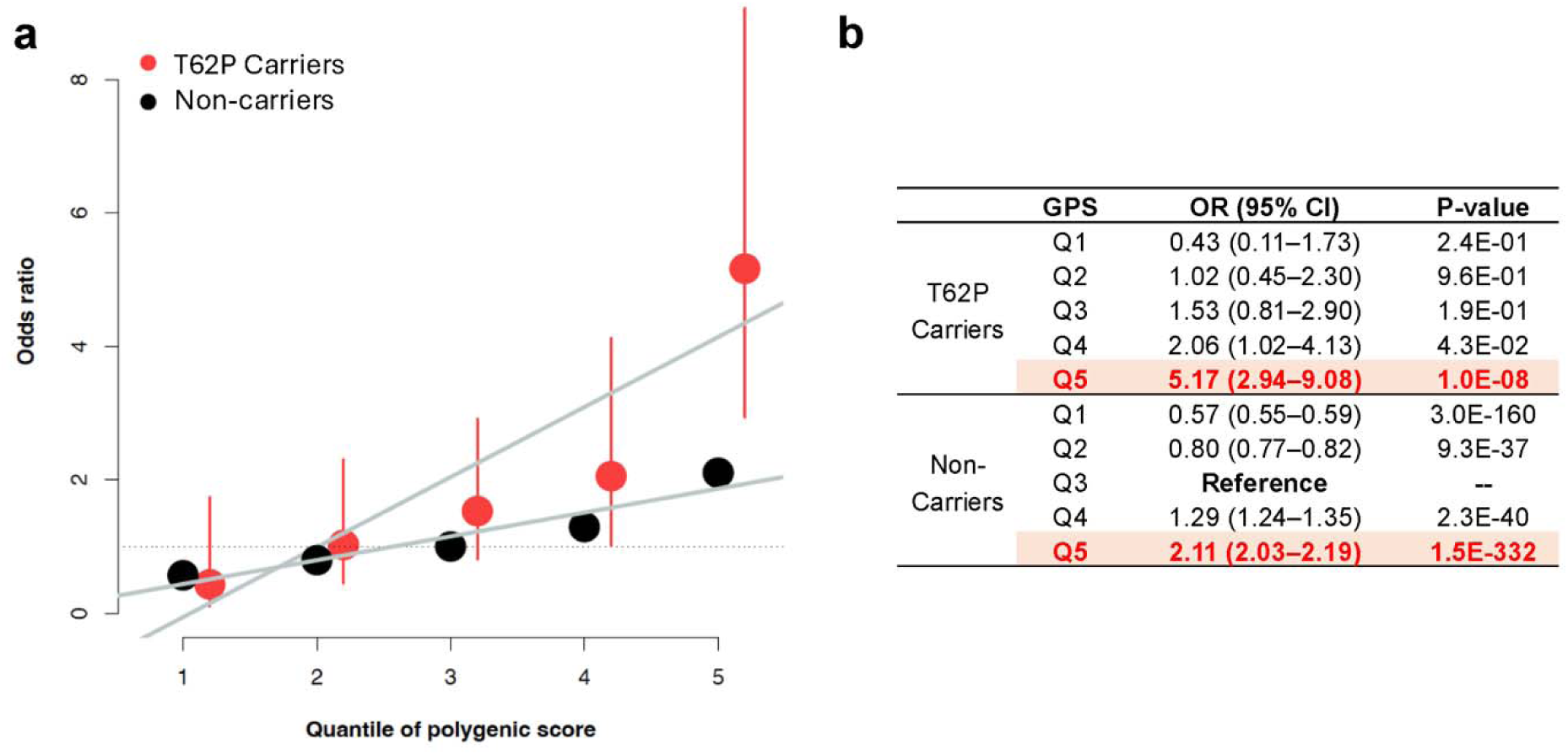
Risk of CKD stratified by *UMOD* T62P carrier status and polygenic risk quintiles: **(a)** Each point represents the odds ratio for CKD relative to the middle GPS quintile among non-carriers. Vertical bars indicate 95% confidence intervals. **(b)** Corresponding effect estimates for each quintile of the GPS. The effect estimates were obtained via meta-analysis of the All of Us and Geisinger cohorts under a fixed-effects model. Individual cohort estimates were adjusted for age, sex, diabetes status, array batch, and principal components of genetic ancestry.

## Discussion

Our study demonstrates that the intermediate-effect *UMOD* T62P variant confers a clinically meaningful but highly variable risk of kidney disease, with the penetrance determined by both age and polygenic background. Across UK Biobank, All of Us, and Geisinger’s MyCode biobanks, T62P carriers had an increased risk of CKD and kidney failure, with penetrance rising substantially with older age. Mechanistically, T62P uromodulin showed impaired maturation, partial ER retention, heightened ER-stress, and increased sensitivity to proteostatic stress compared with wild-type uromodulin, though less severe than canonical ADTKD-causing *UMOD* mutations. Cellular aging further amplified these abnormalities, producing chronic ER-stress/UPR activation, inflammatory remodeling, senescence markers, and tubular injury signals, providing a biologic framework for the observed age-dependent penetrance.

Moreover, polygenic risk was a strong modifier of T62P penetrance, and the GPS effects were markedly larger in T62P carriers than non-carriers. These results produced a striking risk stratification—carriers in the highest GPS quintile had >5-fold higher CKD risk, whereas those in the lowest quintile did not show significantly elevated risk compared to average non-carriers. The observed GPS-by-T62P interaction contrasts with the interplay previously reported for GPS and *APOL1* risk genotypes, for which the risk effects were mostly additive^12,13^. These findings highlight the complexity of the genetic architecture of CKD and may have clinical implications for risk stratification based on genetic testing.

While polygenic scores are increasingly applied to common diseases, their role in modifying the penetrance of monogenic variants is not well established. Our study provides a framework for integrating polygenic scores with monogenic risk in CKD, suggesting that this approach could be particularly meaningful for intermediate frequency and low-penetrance variants. Staged study design and extensive sensitivity analyses assured the robustness of our findings across age groups and biobanks. Moreover, a locus-restricted GPS analysis suggested that regulatory variation near *UMOD* does not fully explain the observed effects, implicating both cis-acting and trans-acting polygenic mechanisms.

Converging evidence suggests that pathogenic *UMOD* variants in ADTKD cause alterations in uromodulin traffic and maturation, causing variable levels of ER stress in tubular cells^8,9,29^. Our previous studies suggested a damaging effect of the intermediate T62P variant on uromodulin maturation, with decreased trafficking to plasma membrane for T62P compared to WT in transiently transfected HEK cells and ER stress in human kidney biopsies^9^. To substantiate the association of T62P variant with kidney injury in a more physiologically relevant system, we now developed stable mIMCD3 cell lines expressing WT, T62P, or the R185S mutant uromodulin causing ADTKD^29^. Compared to WT cells, clones of T62P-expressing cells consistently exhibit a defective uromodulin maturation profile, with impaired ER processing, and increased baseline ER stress. Exposure to DTT confirmed the impact of the T62P variant on defective uromodulin maturation, likely related to disulfide bond formation, and the defective ER homeostasis compared to WT^9^.

Our results also indicate the importance of age for the penetrance of the intermediate effect T62P risk variant. This observation is supported by the fact that T62P carriers with CKD were on average 16 years older than those without CKD (57.5±17.8 vs 41.5±19.4, respectively)^9^. It is also in line with the strong interaction of age with common *UMOD* regulatory variants associated with kidney function by GWAS^16,17^. Increasing the cell passage number induces changes similar to aging^27,28^. Serial passaging is widely used as an *in vitro* model of replicative senescence, capturing the progressive decline in cellular homeostasis including loss of proteostasis, mitochondrial dysfunction, and inflammatory reprogramming^30,31^. In primary renal tubular epithelial models, repeated passaging has been shown to induce senescence-like phenotypes characterized by increased p16 and p21 expression and reduced proliferative capacity, processes linked to kidney aging and CKD^32^. Cellular senescence has also been established as a contributor to kidney disease progression, linking persistent tubular stress responses to chronic inflammation and tissue injury^33,34^.

While immortalized kidney tubular cells are less prone to replicative senescence and are typically driven into senescence through environmental or genetic perturbations, prolonged culture can nevertheless reveal progressive alterations in cellular stress adaptation^35,36^. Here, repeated passaging of mIMCD3 cells expressing T62P led to progressive and coordinated transcriptional remodeling, indicating that continued expression of misfolded uromodulin drives a time-dependent adaptive response. Early transcriptional changes were characterized by activation of UPR and inflammatory pathways, consistent with an acute attempt to restore ER homeostasis. This is supported by induction of stress-associated regulators and enrichment of innate immune and antigen presentation pathways. With continued passaging, however, this response evolved into persistent UPR activation, inflammation, and engagement of signaling pathways linked to cellular remodeling. Sustained expression of inflammatory mediators and late induction of *Wnt5a* and *Axl* suggest a transition toward a chronically adapted state characterized by altered signaling and stress tolerance. In parallel, RNA-seq demonstrated coordinated downregulation of mitochondrial and biosynthetic pathways, indicating a progressive decline in metabolic capacity. Importantly, these transcriptional changes were mirrored at the protein level by accumulation of GRP78, Hsc70, phosphorylated PERK, and eIF2α, together with increased p16, p21, and LCN2 expression, supporting the emergence of a senescence-like phenotype^27,28^.

These findings indicate that T62P and cellular aging exert distinct yet converging effects. Repeated passaging of WT cells also induced modest and transient inflammatory responses, indicating that prolonged culture alone is sufficient to trigger aspects of stress adaptation and replicative remodeling. However, these responses largely resolved over time in WT cells, whereas T62P-expressing cells showed stronger and more persistent activation of ER stress, inflammatory, and remodeling pathways. While T62P imposes a chronic, low-grade proteostatic burden through impaired uromodulin maturation, aging progressively reduces the cellular capacity to maintain proteostasis, alters metabolic fitness, and promotes a pro-inflammatory state. Together, these processes amplify ER stress and promote maladaptive responses over time. Ultimately, this interplay may increase the susceptibility of tubular cells to injury and contribute to progressive kidney damage^37^. Consistent with these results, we observed a strong age-dependent penetrance of T62P kidney disease manifestations in our cross-biobank analyses. Notably, these allelic effects could be amplified by the exposure to kidney stressors over time, including ischemic or toxic damage, as already suggested for the effect of common *UMOD* promoter variants on the risk of CKD^38^.

Several limitations of our study should be noted. First, the biobank analyses have a pragmatic cross-sectional design necessitated by the sample size requirements for testing interactions with low frequency alleles. We recognize that prospective studies of T62P carriers are still needed to refine the role of GPS in predicting incident kidney failure. Second, there are inherent limitations of biobank-linked EHR data, such as non-random missingness or potential for disease miscoding. To mitigate these issues, we relied on the validated e-phenotype for diagnosing and staging CKD based on laboratory data rather than billing codes alone. More granular phenotypic data on the subtypes of CKD, such as kidney biopsy diagnoses, would further enrich our analyses, but such data are presently not available for biobanks. Third, our *in vitro* studies are limited by the use of mIMCD cell lines. While no human cell lines producing significant amounts of endogenous uromodulin are currently available, mIMCD cell lines have been used extensively to study the effects of canonical *UMOD* variants^29^. Lastly, we remain underpowered to test whether the GPS has an effect on the disease course in carriers of canonical pathogenic missense ADTKD variants, mainly because these variants are typically ultra-rare with insufficient numbers of carriers even across very large biobanks. Further studies of the GPS in the collections of ADTKD pedigrees segregating pathogenic *UMOD* variants would be needed to address this limitation.

In summary, *UMOD* T62P illustrates a complex genetic model in which kidney disease risk is shaped by the combined effects of variant-specific proteotoxicity, age-related loss of tubular proteostatic capacity, and genome-wide polygenic susceptibility. The T62P variant imposes a chronic, partially compensated defect in uromodulin maturation and ER trafficking. With aging, diminished adaptive reserve and sustained UPR activation may shift this defect toward tubular injury and CKD. The effect modification of T62P-associated risk by GPS indicates that polygenic factors can shape the penetrance of intermediate-effect risk alleles. These findings support a contextual model of inherited kidney disease risk, in which variant interpretation and clinical risk prediction incorporate not only the causal allele, but also polygenic background and other biological modifiers.

## Data Availability

The UKBB genotype and phenotype data are available through the UKBB web portal at https://www.ukbiobank.ac.uk/. Researchers wishing to access these resources must register with the UKBB. Similarly, the AoU genotype, WGS, and phenotype data are accessible via the AoU Researcher Workbench at https://www.researchallofus.org/data-tools/workbench/. Researchers interested in these data must complete registration with the AoU study. Both biobanks also require institutional data use agreements as part of the registration process. Due to consent restrictions, MyCode data is not accessible on the cloud platform, but relevant data can be made available upon reasonable request to the corresponding authors.

https://www.researchallofus.org/data-tools/workbench/

## Acknowledgements

We are grateful to all participants from the UKBB, AoU, and MyCode projects for contributing their data and biological samples that enabled this study, and to John A. Sayer (Newcastle University, Newcastle upon Tyne, UK) and Peter J. Conlon (Beaumont Hospital, Dublin, Ireland) for contributing samples and discussions. The data from the MyCode project reported here have been supplied by the United States Renal Data System (USRDS). The interpretation and reporting of these data are the responsibility of the author(s) and should not be seen as an official policy or interpretation of the U.S. government. This work was funded by the National Institute for Diabetes and Digestive Diseases (NIDDK) grants R01-DK144793, R03DK144285, and K25-DK128563, National Human Genome Research Institute (NHGRI) Electronic Medical Records and Genomics-IV (eMERGE-IV) grant 5U01-HG008680, National Library of Medicine (NLM) grant R01LM013061, and National Center for Advancing Translational Sciences (NCATS) grant UL1TR001873. The research on UK Biobank data has been conducted using the UK Biobank Resource under Application Number 41849. The All of Us Research Program is supported by the NIH Office of the Director through the following grants: Regional Medical Centers: 1OT2OD026549; 1OT2OD026554; 1OT2OD026557; 1OT2OD026556; 1OT2OD026550; 1OT2OD 026552; 1OT2OD026553; 1OT2OD026548; 1OT2OD026551; 1OT2OD026555; IAA# AOD16037; Federally Qualified Health Centers: HHSN 263201600085U; Data and Research Center: 5U2COD023196; Biobank: 1U24OD023121; The Participant Center: U24OD023176; Participant Technology Systems Center: 1U24OD023163; Communications and Engagement: 3OT2OD023205; 3OT2OD023206; and Community Partners: 1OT2OD025277; 3OT2OD025315; 1OT2OD025337; 1OT2OD025276. M.M and O.D. are supported by the European Union’s Horizon 2020 research and innovation program under the Marie Skłodowska-Curie grant (agreement N° 860977). O.D. and E.O. are supported by the European Reference Network for Rare Kidney Diseases (project N° 739532) and the ADTKD-Net project of the ERA-NET + EJP 2023 (grant 222473). O.D. is supported by the Swiss National Science Foundation (grant 10.003.608); the University Research Priority Program (URPP) ITINERARE at the University of Zurich; and the Saint-Luc Foundation (UCLouvain, Brussels).

## Contributions

Project conceptualization: K.K., O.D., A.K., E.O.; statistical methods and analysis plan for biobank data: A.K., K.K.; electronic CKD phenotyping: N.S.; variant filtering: A.K., K.K., M.V.P.G., O.D., E.O., H.M.; MyCode replication analysis: A.C., I.D., T.M.; cell culture studies: E.O., A.G., M.M., O.D.; manuscript draft: A.K., K.K., O.D.; overall supervision: K.K.

## Supplementary Information

### Biobank Datasets

#### UK Biobank (UKBB)

The design and composition of this dataset have been previously described in detail^1^. We accessed and analyzed the most recent data via the UKBB Research Analysis Platform (RAP) on DNAnexus. To extract UMOD variant information, we used exome sequence data for 469,835 participants as previously described^2,3^. The GPS for CKD was calculated based on the imputed microarray data for those individuals who had exome sequencing available. We implemented electronic phenotyping for CKD in this cohort, which resulted in the final number of 10767 cases (CKD stage 3 or above) and 282647 controls included in the downstream statistical analyses.

#### All-of-US (AoU)

The All-of-Us research program has been described in detail^4^. We used the second data release that included N=312,944 participants. All analyses were performed using AoU Workbench on Google Cloud. To extract UMOD variant information, we used genome sequence data. To calculate the GPS, we used imputed genotype data as described previously^5^. Finally, we implemented our electronic CKD phenotype algorithm to define 26521 cases (CKD stage 3 or above) and 80753 controls for downstream statistical analyses.

#### MyCode

The Geisinger MyCode Community Health Initiative has been described previously in detail^6^. Briefly, MyCode is an integrated research biobank that links electronic health record (EHR) data with genomic data obtained from participants across the Geisinger Health System in Pennsylvania. To extract *UMOD* variant information, we used exome sequencing data from 167,892 participants generated through the DiscovEHR collaboration with Regeneron Genetics Center, and quality-controlled according to previously published protocols^6^. Genotype imputation was performed using microarray data available for the same individuals, allowing calculation of the GPS for CKD in participants with both imputed array data and exome data. We applied an electronic CKD phenotype algorithm to the linked EHR data within MyCode, defining CKD stage 3 or above based on laboratory measurements and diagnostic codes. After application of all inclusion/exclusion criteria, the final dataset consisted of 27299 cases (CKD stage 3 or above) and 65979 controls for downstream statistical analyses. For ESKD, the MyCode cohort used linkage to the USRDS database^7^, the national data registry that collects, analyzes, and distributes information about the ESKD population in the U.S, and included 1773 cases and 166119 controls for downstream statistical analyses.

---

**Supplementary Figure 1.**
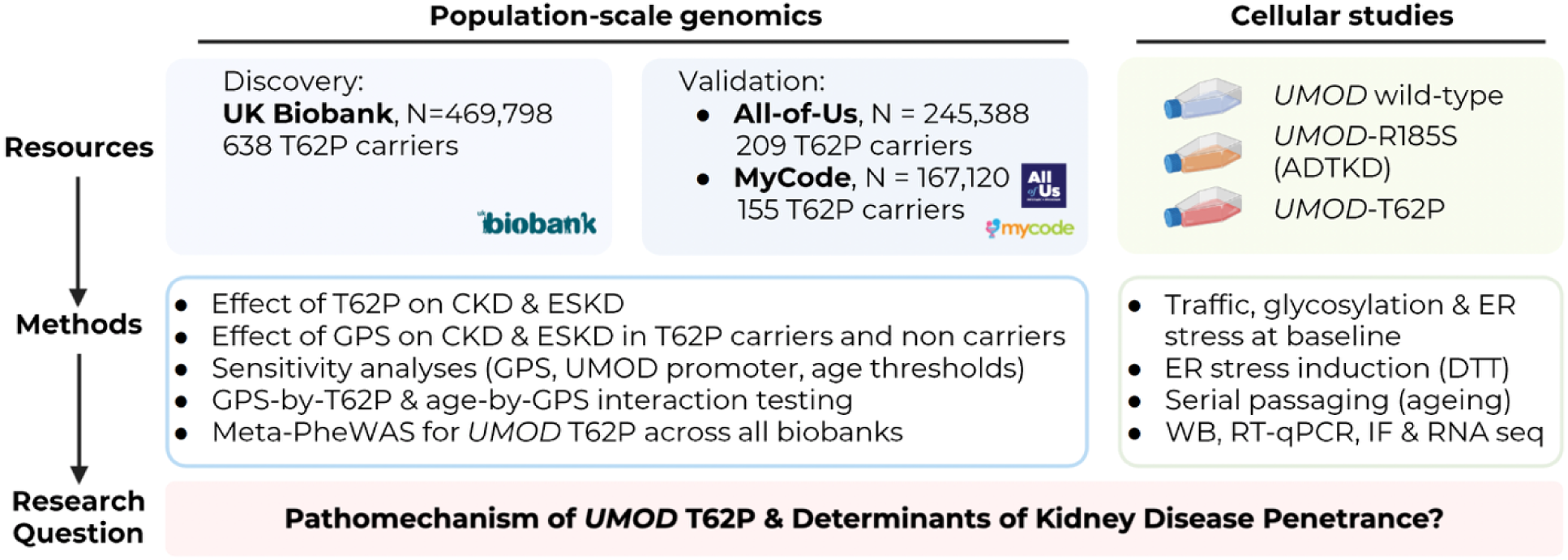
Study workflow. Flow of the population and cellular studies investigating the determinants of the penetrance of the intermediate *UMOD* T62P variant.

**Supplementary Figure 2.**
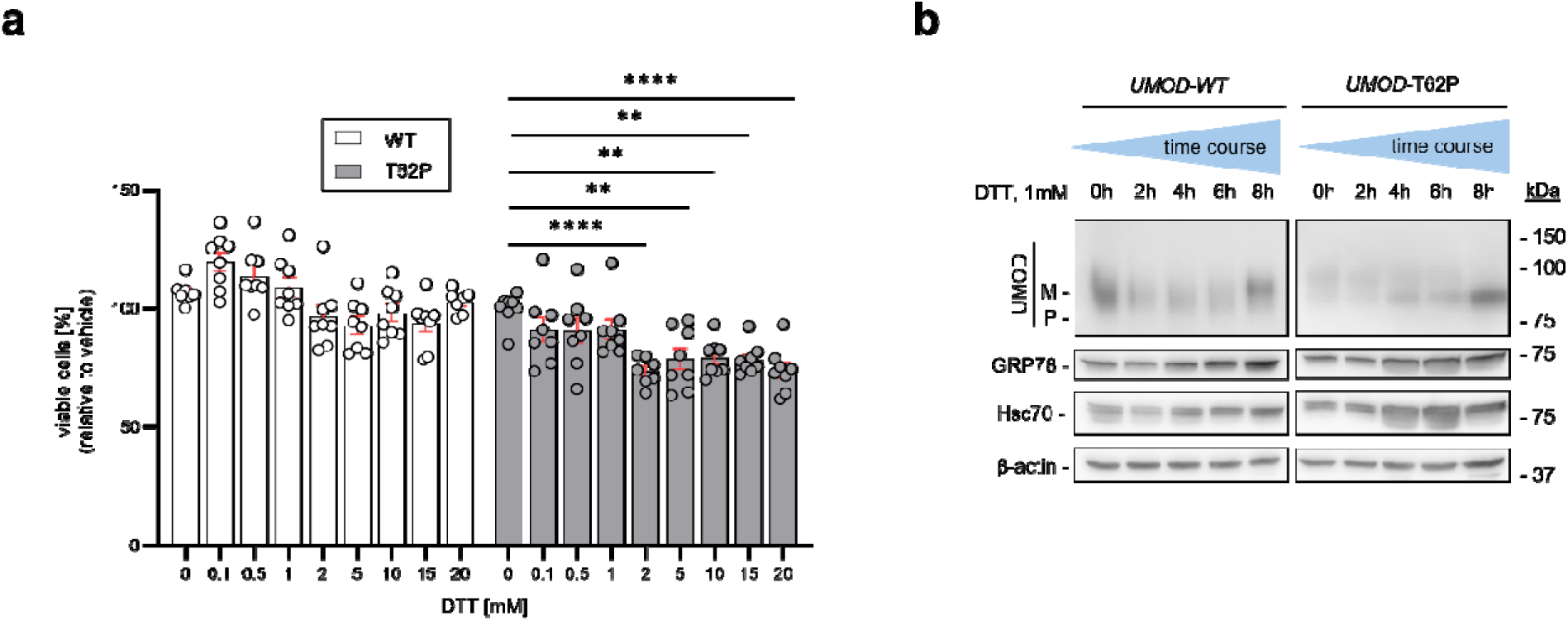
Dose- and time course-dependent effects of DTT treatment on cells expressing *UMOD* variants. (**a**) MTT cell viability assay of *UMOD*-wild-type (WT) and T62P expressing cells, exposed to indicated concentrations of DTT for 8h. Bars represent mean ± SEM. Ordinary one-way ANOVA followed by Dunnett’s multiple comparisons test. *P<0.05, **P<0.01, ***P<0.001, ****P<0.0001. (**b**) Immunoblot of protein lysates from WT- or T62P-expressing cells following time-course treatment with 1mM DTT for the indicated duration, probed for UMOD, GRP78 and Hsc70 with β-actin serving as a loading control.

**Supplementary Figure 3.**
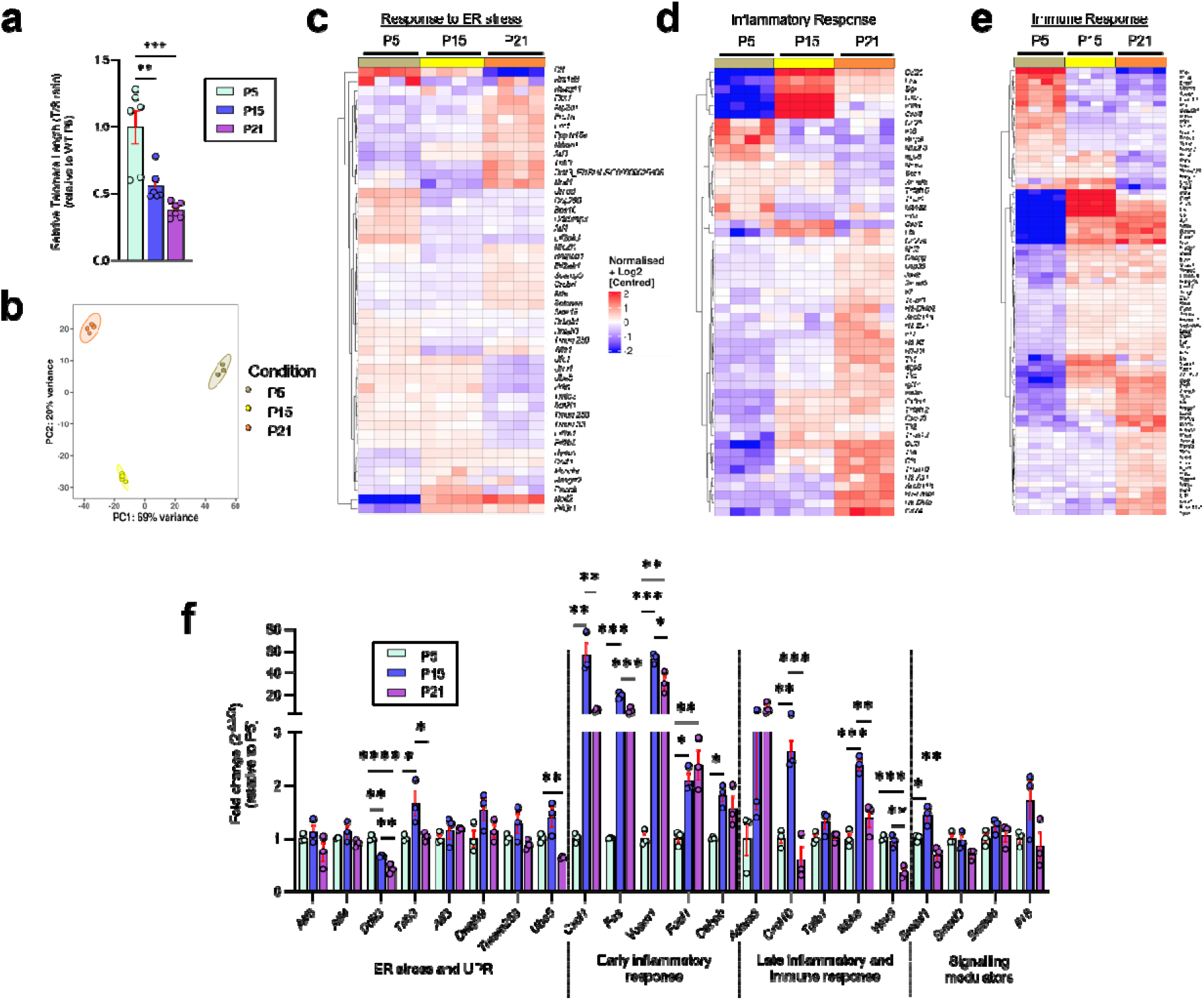
Effects of increasing passage numbers on telomere length, response to ER stress, inflammatory and immune response in T62P and wild-type UMOD expressing cells. **(a)** Relative telomer length of *UMOD*-WT expressing mIMCD3 cells harvested at passages 5 (P5), 15 (P15), and 21 (P21) (n=6 biological replicates). Bars represent mean ± SEM. Ordinary one-way ANOVA followed by Tukey’s post hoc test. *P<0.05, **P<0.01, ***P<0.001, ****P<0.0001. **(b)** Principal component analysis (PCA) of RNA-sequencing data from T62P-expressing mIMCD3 cells harvested at passages 5 (P5), 15 (P15), and 21 (P21). PC1 and PC2 explain 69% and 20% of the total variance, respectively. **(c-e)** Heatmaps of differently expressed genes (DEGs) in selected Gene Ontology (GO) pathways across passages (P5, P15, P21) related to **(c)** response to endoplasmic reticulum stress, **(d)** immune response, and **(e)** inflammatory response. Data are shown as log_2_-normalized counts, centered by gene. **(f)** RT-qPCR validation of ER-stress and unfolded protein response (UPR), inflammatory and immune response, and signaling modulator genes in *UMOD*-WT-expressing cells at passages 5, 15 and 21 (n=3 biological replicates). Bars represent mean ± SEM. Ordinary one-way ANOVA followed by Tukey’s post hoc test. *P<0.05, **P<0.01, ***P<0.001, ****P<0.0001.

**Supplementary Table 1.** The performance of GPS in a re-optimization analysis using 70% of the UKBB after excluding all carriers of the T62P variant. Performance was assessed using logistic regression models with the GPS as the predictor and CKD case-control status as the outcome. Reported metrics include odds ratio (OR) per standard deviation (SD) of each GPS and corresponding P-values. The models were adjusted for age, sex, diabetes status, genotyping batch, and genetic ancestry. The adjusted area under the receiver operating characteristic curve (AUC) reflects performance including covariates, while the crude AUC reflects performance of the GPS alone. Nagelkerke’s pseudo-R² was used to estimate the variance in CKD status explained by the GPS, independent of covariates. For P+T models, *r²* indicates the LD pruning threshold; for LDpred, ρ (rho) denotes the fraction of causal variants assumed. The best-performing GPS is highlighted in bold red.

| Method | Parameter | N variants | OR* per SD of GPS | P-value | AUC (Adjusted*) | AUC (Crude) | Nagelkerke $R^2$ |
| --- | --- | --- | --- | --- | --- | --- | --- |
| P+T | P=1E-01 | 89,880 | 1.85 | P<1.00E-300 | 0.8382 | 0.6507 | 0.0439 |
| P+T | P=1E-02 | 21,764 | 1.84 | P<1.00E-300 | 0.8388 | 0.6511 | 0.044 |
| P+T | P=1E-03 | 7,486 | 1.77 | P<1.00E-300 | 0.8361 | 0.6415 | 0.0389 |
| P+T | P=1E-04 | 3,598 | 1.76 | P<1.00E-300 | 0.8356 | 0.6391 | 0.038 |
| P+T | P=1E-05 | 2,111 | 1.74 | P<1.00E-300 | 0.835 | 0.6374 | 0.0367 |
| P+T | P=1E-06 | 1,407 | 1.74 | P<1.00E-300 | 0.8349 | 0.6369 | 0.0363 |
| P+T | P=1E-07 | 1,028 | 1.74 | P<1.00E-300 | 0.8345 | 0.6366 | 0.0361 |
| P+T | P=1E-08 | 753 | 1.72 | P<1.00E-300 | 0.8336 | 0.6341 | 0.0345 |
| P+T | P=3.0E-02 | 41,426 | 1.85 | P<1.00E-300 | 0.8388 | 0.6522 | 0.0446 |
| P+T | P=3.0E-03 | 11,918 | 1.81 | P<1.00E-300 | 0.8374 | 0.6464 | 0.0415 |
| P+T | P=3.0E-04 | 4,971 | 1.77 | P<1.00E-300 | 0.8362 | 0.6412 | 0.039 |
| P+T | P=3.0E-05 | 2,675 | 1.75 | P<1.00E-300 | 0.8352 | 0.6378 | 0.0373 |
| LDpred | $\rho=1.0E+00$ | 5,440,627 | 1.83 | P<1.00E-300 | 0.8376 | 0.6505 | 0.043 |
| LDpred | $\rho=1.0E-01$ | 5,440,627 | 1.17 | 1.51E-31 | 0.8153 | 0.5378 | 0.0027 |
| LDpred | $\rho=1.0E-02$ | 5,440,627 | 1.16 | 3.46E-30 | 0.8149 | 0.5394 | 0.0026 |
| LDpred | $\rho=1.0E-03$ | 5,440,627 | 1.16 | 4.64E-29 | 0.8149 | 0.5385 | 0.0025 |
| LDpred | $\rho=3.0E-01$ | 5,440,627 | 1.64 | 1.11E-307 | 0.8296 | 0.6242 | 0.0286 |
| LDpred | $\rho=3.0E-02$ | 5,440,627 | 1.17 | 3.74E-32 | 0.8152 | 0.5376 | 0.0027 |
| LDpred | $\rho=3.0E-03$ | 5,440,627 | 1.16 | 9.38E-30 | 0.815 | 0.539 | 0.0025 |

\*Adjusted for age, sex, diabetes, the first four principal ancestry components, and genotyping batch.

**Supplementary Table 2.** Sensitivity analysis after exclusion of *UMOD* locus from the genome-wide polygenic score (GPS). UKBB-based associations between the GPS and CKD risk among carriers of the *UMOD* T62P variant, before and after exclusion of variants located within ±200 kb of the *UMOD* gene. Effect estimates for each predictor (T62P variant; GPS among T62P carriers; and GPS among T62P non-carriers) are adjusted for age, sex, diabetes status, array batch (if applicable), and genetic ancestry.

|  | **GPS in T62P Carriers** | **GPS in Non-Carriers** |
| --- | --- | --- |
|  | **OR per SD (95%CI), P-value** | **OR per SD (95%CI), P-value** |
| **GPS with *UMOD* locus** | 4.68 (3.03 – 7.11), P=8.9E-05 | 1.83 (1.81 – 1.85), P<E-300 |
| **GPS without *UMOD* locus** | 4.66 (2.15 – 10.1), P=8.9E-05 | 1.83 (1.79 – 1.87), P<E-300 |

**Supplementary Table 3.**
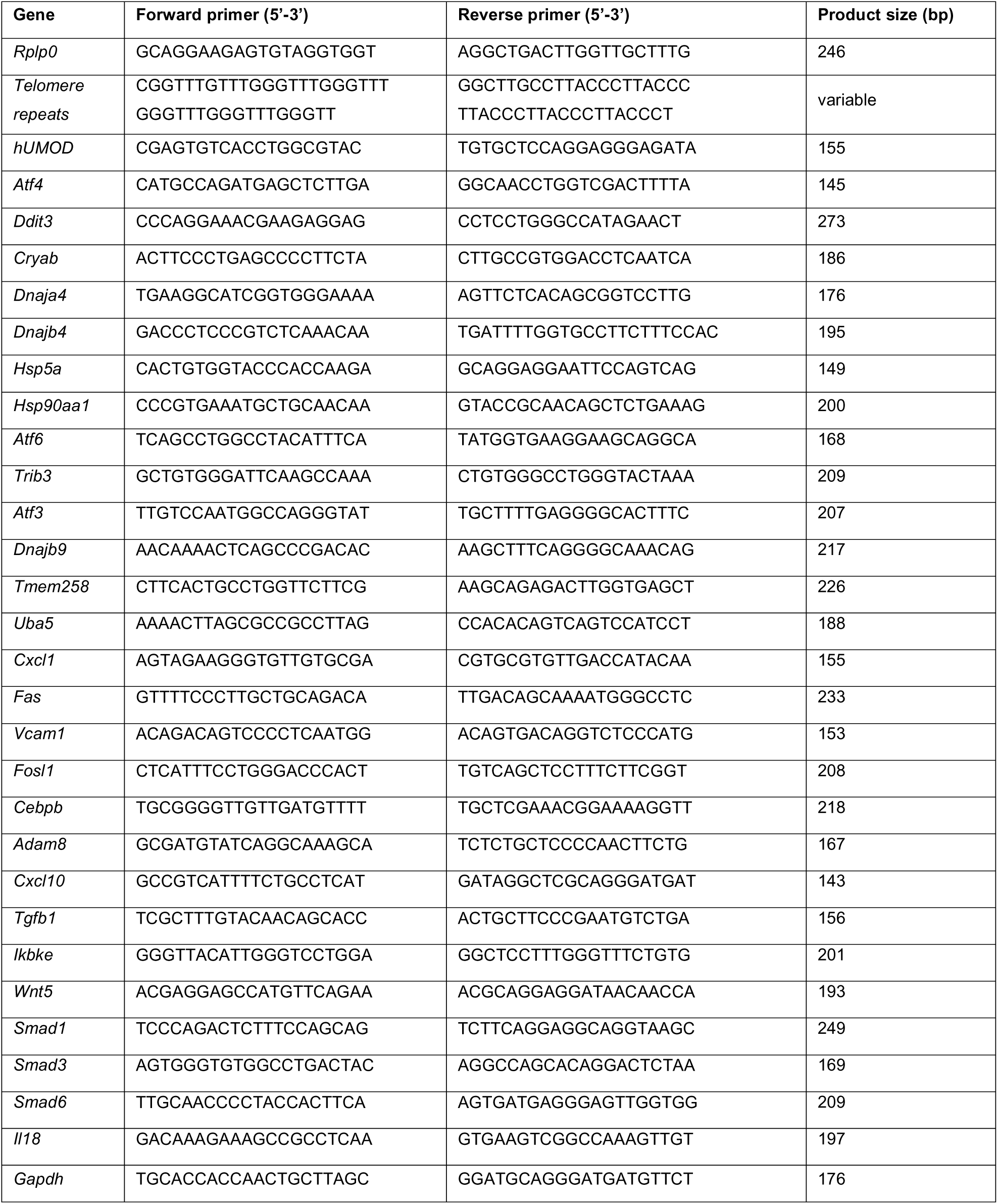
List of primers used for RT-qPCR analysis.

**Supplementary Table 4.** List of primary antibodies.

| Target antigen | Host species | Dilution | Cat # | Source |
| --- | --- | --- | --- | --- |
| hUromodulin | Sheep | WB: 1:500 | PA546959 | Invitrogen |
|  |  | IF: 1:600 |  |  |
| GRP78/BiP | Rabbit | WB 1:1000 | ab21685 | Abcam |
|  |  | IF 1:400 |  |  |
| Hsc70 | Rabbit | WB 1:1000 | ab51052 | Abcam |
| Lipocalin-2 (LCN2) | Goat | WB 1:500 | AF1857 | R&D Systems |
| eIF2α (D7D3) | Rabbit | WB: 1:500 | #5324 | Cell Signaling |
| phospho-eIF2α<br>(Ser51) (D9G8) | Rabbit | WB: 1:500 | #3398 | Cell Signaling |
| ATF-4 (D4B8) | Rabbit | WB 1:500 | #11815 | Cell Signaling |
| PERK (C33E10) | Rabbit | WB: 1:500 | #3192 | Cell Signaling |
| Phospho-PERK | Rabbit | WB: 1:500 | MA5-15033 | Invitrogen |
| p21 (EPR18021) | Rabbit | WB: 1:1000 | ab188224 | Abcam |
| CDKN2A/p16INK4a<br>(EPR20418) | Rabbit | WB: 1:1000 | ab211542 | Abcam |
| β-Actin | Mouse | WB 1:10000 | A5441 | Sigma-Aldrich |
| Calnexin | Rabbit | IF: 1:400 | C4731 | Sigma-Aldrich |

**Supplementary Table 5.** CKD risk estimates stratified by T62P carrier status and quantiles of GPS risk. Each Odds Ratio (OR) was calculated in reference to the middle GPS quantile for non-carriers; all effect estimates were adjusted for age, sex, diabetes, array batch, and genetic ancestry; data from individual cohorts w*ere* pooled using fixed effects meta-analysis.

| Biobank | Quintile | T62P carriers | T62P non-carriers |
| --- | --- | --- | --- |
|  |  | OR (95% CI), P | OR (95% CI) |
| UKBB<br>(Discovery) | Quintile 1 | 0.00 (0.00–>100), 9.39E-01 | 0.49 (0.45–0.53), 3.25E-63 |
|  | Quintile 2 | 0.74 (0.18–3.10), 6.79E-01 | 0.74 (0.68–0.80), 2.10E-15 |
|  | Quintile 3 | 2.60 (1.09–6.20), 3.19E-02 | **Reference** |
|  | Quintile 4 | 2.92 (1.22–6.97), 1.60E-02 | 1.40 (1.32–1.50), 5.96E-24 |
|  | **Quintile 5** | **6.96 (3.56–13.6), 1.50E-08** | **2.60 (2.44–2.76), 1.42E-208** |
| AoU<br>(Validation 1) | Quintile 1 | 0.26 (0.03–2.20), 2.15E-01 | 0.66 (0.62–0.71), 1.09E-30 |
|  | Quintile 2 | 2.24 (0.62–8.16), 2.21E-01 | 0.86 (0.80–0.92), 1.13E-05 |
|  | Quintile 3 | 0.75 (0.19–2.94), 6.76E-01 | **Reference** |
|  | Quintile 4 | 1.17 (0.29–4.73), 8.21E-01 | 1.15 (1.07–1.23), 6.41E-05 |
|  | **Quintile 5** | **1.57 (0.45–5.47), 4.77E-01** | **1.64 (1.53–1.75), 1.32E-45** |
| MyCode<br>(Validation 2) | Quintile 1 | 0.63 (0.10–3.96), 6.10E-01 | 0.55 (0.51–0.59), 0 |
|  | Quintile 2 | 0.50 (0.11–2.27), 3.50E-01 | 0.79 (0.75–0.83), 0 |
|  | Quintile 3 | 0.89 (0.24–3.29), 8.60E-01 | **Reference** |
|  | Quintile 4 | 0.97 (0.12–7.86), 9.80E-01 | 1.33 (1.25–1.42), 0 |
|  | **Quintile 5** | **7.33 (1.16–46.3), 3.00E-02** | **2.09 (1.96–2.23), 0** |
| Validation Meta<br>(AoU +<br>MyCode) | Quintile 1 | 0.43 (0.10–1.73), 2.38E-01 | 0.59 (0.57–0.63), P=2.37E-102 |
|  | Quintile 2 | 1.19 (0.44–3.17), 7.22E-01 | 0.812 (0.77–0.84), P=4.8E-24 |
|  | Quintile 3 | 0.81 (0.31–2.10), 6.78E-10 | **Reference** |
|  | Quintile 4 | 1.10 (0.34–3.53), 8.63E-01 | 1.241 (1.17–1.30), P=4.523E-20 |
|  | **Quintile 5** | **2.54 (0.90–7.16), 7.59E-02** | **1.854 (1.76–1.95), P=1.34E-142** |
| Combined Meta<br>(Discovery +<br>Validation) | Quintile 1 | 0.43 (0.11–1.73), 2.37E-01 | 0.57 (0.55–0.59), 2.97E-160 |
|  | Quintile 2 | 1.02 (0.45–2.30), 9.56E-01 | 0.80 (0.77–0.82), 9.29E-37 |
|  | Quintile 3 | 1.53 (0.81–2.90), 1.93E-01 | **Reference** |
|  | Quintile 4 | 2.06 (1.02–4.13), 4.28E-02 | 1.29 (1.24–1.35), 2.31E-40 |
|  | **Quintile 5** | **5.17 (2.94–9.08), 1.04E-08** | **2.11 (2.03–2.19), 1.53E-332** |

## Notes

### Competing Interest Statement

The authors have declared no competing interest.

